# Resource Considerations for Acute Ischemic Stroke Intervention in Nonagenarians

**DOI:** 10.1101/2024.07.24.24310904

**Authors:** Sani Gandhi, Kimberly Hollender, Florence Chukwuneke, Deviyani Mehta, Roger Cheng

## Abstract

**Background:** While there is evidence to support the viability of intravenous thrombolysis (IVT) and mechanical thrombectomy (MT) as treatments for acute ischemic stroke (AIS) even in the very elderly, there are practical considerations to be made regarding triage and resource allocation, particularly in a hub-and-spoke system of stroke intervention.

**Methods:** This is a cross-sectional study using stroke quality registry data from 2017-2021 at a comprehensive stroke center serving as the primary hub for an associated healthcare system. We identified patients aged 90 or above who received acute stroke treatment with IVT and/or MT. NIHSS, modified Rankin Score (mRS), transfer status, length of stay (LOS), and discharge disposition were examined.

**Results:** Out of 268 total nonagenarians admitted for AIS, 60 received an acute intervention (37 IVT, 14 MT, 9 both). All MT attempts resulted in successful reperfusion (>TICI2b). Median initial and discharge mRS were 2 and 5 respectively; only 3.3% were discharged home, with 46.7% of patients either deceased or discharged to hospice. Median LOS was 5 days (range 0-77), but prolonged LOS was common, with 11 patients having LOS >14 days. Comparing the groups who received MT vs. IVT alone, the median initial NIHSS was 17 vs. 15 (p=0.23); the MT group had better baseline mRS (1 vs 3, p=0.005), but despite this, there were no significant differences in mortality (57% vs 41%) or discharge mRS (5 vs. 5). Median LOS was not significantly different (6 vs. 5 days). Mortality in MT was 75% for transfers vs. 53% for local arrivals (p=0.36).

**Conclusion:** Despite safe and technically successful treatment, outcomes were poor overall in the nonagenarian population after both MT +/- IVT, with very high morbidity and mortality.

## INTRODUCTION

Advances in stroke treatments over the past decade, such as mechanical thrombectomy (MT) for large vessel occlusion (LVO) and widespread availability of intravenous thrombolysis (IVT) have both improved clinical outcomes and broadened the indications for acute intervention. However, evidence is still inconsistent for treatment of the extremely elderly, particularly patients above age 90, a population that was specifically excluded from major MT trials ^[1][2][3]^. Functional outcomes with thrombectomy have been reported as being up to five times worse in nonagenarians despite evidence demonstrating safety and procedural efficacy ^[4][5][6][7][8]^. In a hub-and-spoke system of stroke intervention, there are also other practical considerations such as resource allocation and whether long-distance transfer is the best patient-centered choice for what frequently becomes end-of-life care. In this retrospective study, we explore the outcomes of IVT and MT in a cohort of nonagenarians presenting with AIS.

## METHODS

### Population

This study is a cross-sectional study of nonagenarians presenting with AIS who received treatment with IVT and/or MT from 2017-2021 at a comprehensive stroke center serving as the primary hub for its associated healthcare system. Deidentified source data was used, comprising of measures and outcomes reported to the state and national stroke registries.

### Interventional Parameters

Patients all underwent imaging with some combination of CT, CT angiography, CT perfusion, and/or MRI. IVT was administered based on disabling stroke deficits and presentation within the <4.5-hour window from the last known well (LKW). Patients selected for MT all had LVO and either met standard criteria for intervention due to being within 6 hours from LKW, or had a mismatch between CBF<30% and Tmax>6 seconds of >1.8 on CT perfusion, per DAWN and DEFUSE-3 criteria^[1][2]^. All MT attempts in the study population resulted in successful recanalization (defined as TICI 2b or above).

### Study Endpoints

The primary outcome was based on the modified Rankin score (mRS) at baseline and hospital discharge. Secondary outcomes included the NIH stroke scale (NIHSS) at discharge and discharge disposition. Other variables that were evaluated include transfer status and hospital length of stay.

### Statistical Analysis

Fisher exact test, Wilcoxon Rank Sum test, and T-test were used for comparisons. Clinical significance was set at p<0.05 and all results were two-sided. Statistical analysis was performed utilizing OriginLab software.

## OUTCOMES

Out of 268 nonagenarian patients who presented from 2017 to 2021 with AIS, 60 patients received intervention. The median age of this group was 93; 75% were female and 81.6% had a white ethnic background. Out of the sample, 37 patients received IVT alone with r-tPA; 14 patients with a LVO received MT alone, and 9 received both interventions (Table 1). The overall symptomatic hemorrhagic transformation rate (sICH) was 3.8%.

**Table 1:**
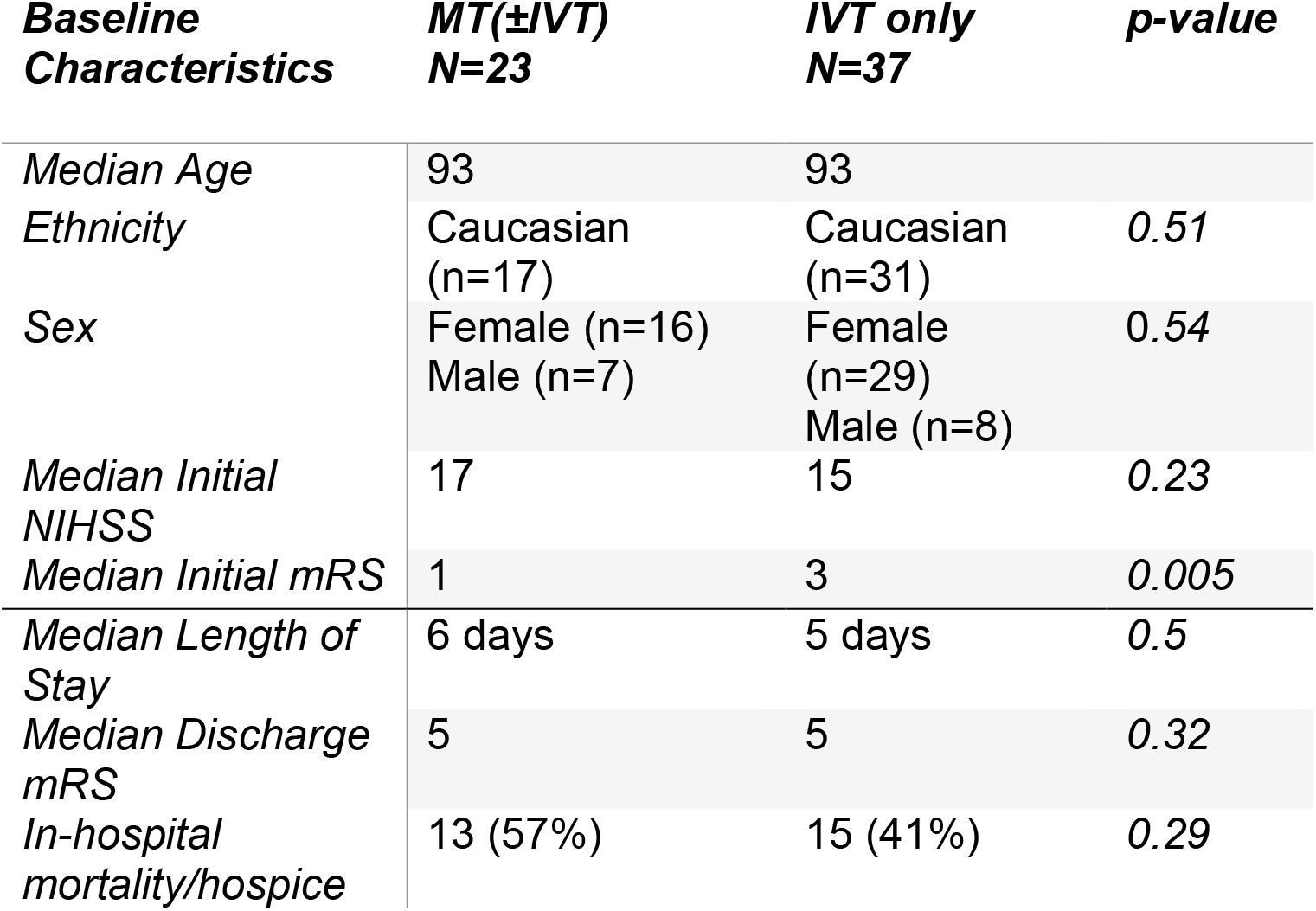
Study population baseline characteristics and outcomes.

| **Baseline Characteristics** | **MT(±IVT)**<br>**N=23** | **IVT only**<br>**N=37** | **p-value** |
| --- | --- | --- | --- |
| *Median Age* | 93 | 93 |  |
| *Ethnicity* | Caucasian<br>(n=17) | Caucasian<br>(n=31) | 0.51 |
| *Sex* | Female (n=16)<br>Male (n=7) | Female<br>(n=29)<br>Male (n=8) | 0.54 |
| *Median Initial NIHSS* | 17 | 15 | 0.23 |
| *Median Initial mRS* | 1 | 3 | 0.005 |
| *Median Length of Stay* | 6 days | 5 days | 0.5 |
| *Median Discharge mRS* | 5 | 5 | 0.32 |
| *In-hospital mortality/hospice* | 13 (57%) | 15 (41%) | 0.29 |

The 23 patients who received MT +/- IVT arrived to the hospital with a median initial NIHSS of 17 (*p=0*.*23)* and had a better baseline median mRS of 1 when compared to the 37 patients who received IVT alone, with median initial NIHSS of 15 and median initial mRS of 3 (*p=0*.*005)*. There was no significant difference in discharge mRS for both groups, with a median score of 5 (*p=0*.*32)*. The median length of stay was 5 days for both intervention groups (range 0-77, *p=0*.*5*); 11 patients (18%) had hospital stays longer than 14 days (Table 1).

More strikingly, of this 60-patient cohort, only 2 patients (3.3%) were discharged to home with mRS<3. Conversely, 28 patients (46.7%) either died in-hospital or were transitioned to hospice. The remaining 50% (n=30) of the patients were discharged to acute or subacute rehabilitation, all with mRS>3 (Figure 1).

**Figure 1:**
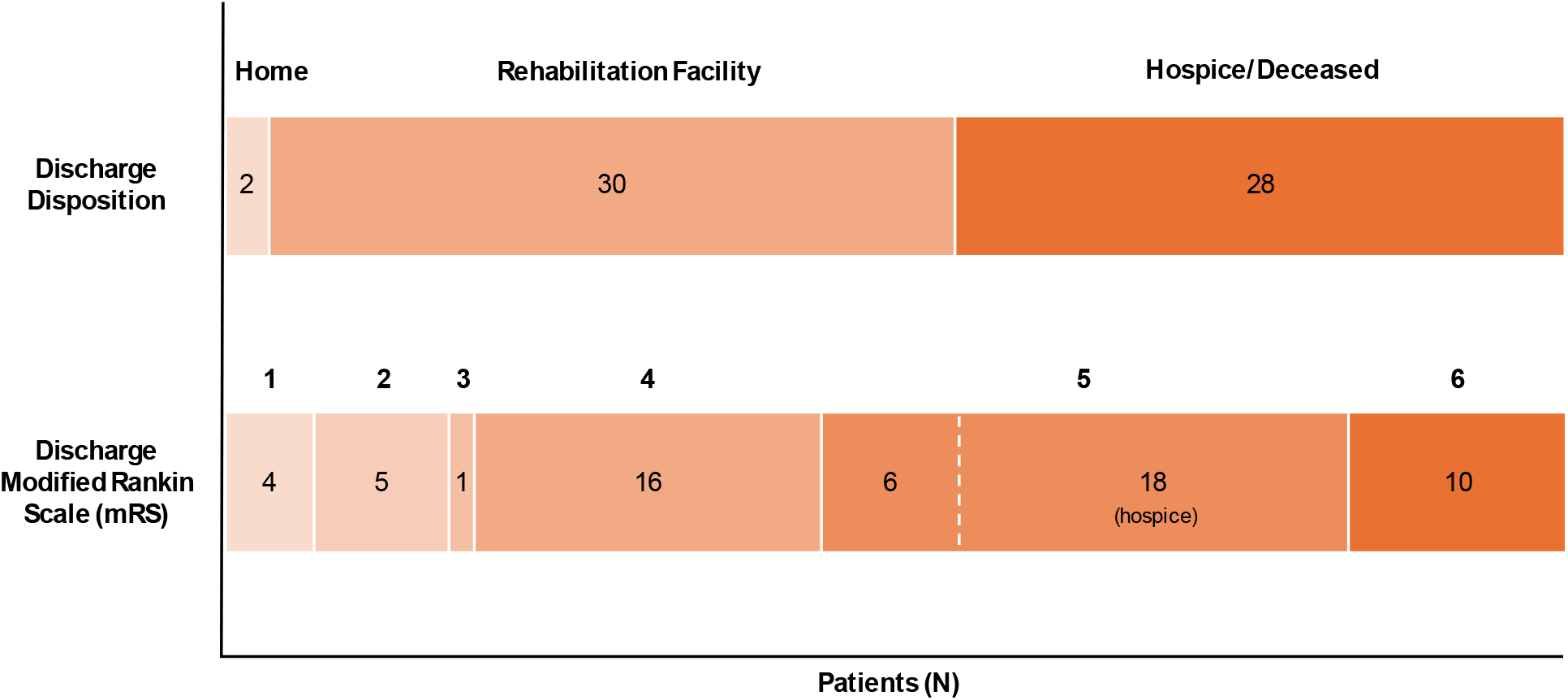
Overall discharge disposition and modified Rankin scale. Overall discharge disposition as well as the modified Rankin scale (mRS) on discharge for the nonagenarian study population receiving acute stroke treatment (60 patients total).

When comparing mortality between interventions, patients who underwent MT +/- IVT had a higher mortality rate of 57% as compared to patients who received IVT alone (41%), despite presenting with a better baseline mRS (1 vs 3). Out of the 23 patients who underwent thrombectomy, 8 arrived as an inter-facility transfer; of these patients, mortality was even higher at 75%, compared to 53% for primary arrivals at our facility, though this was not a significant difference (*p=0*.*36)* (Figure 2).

**Figure 2:**
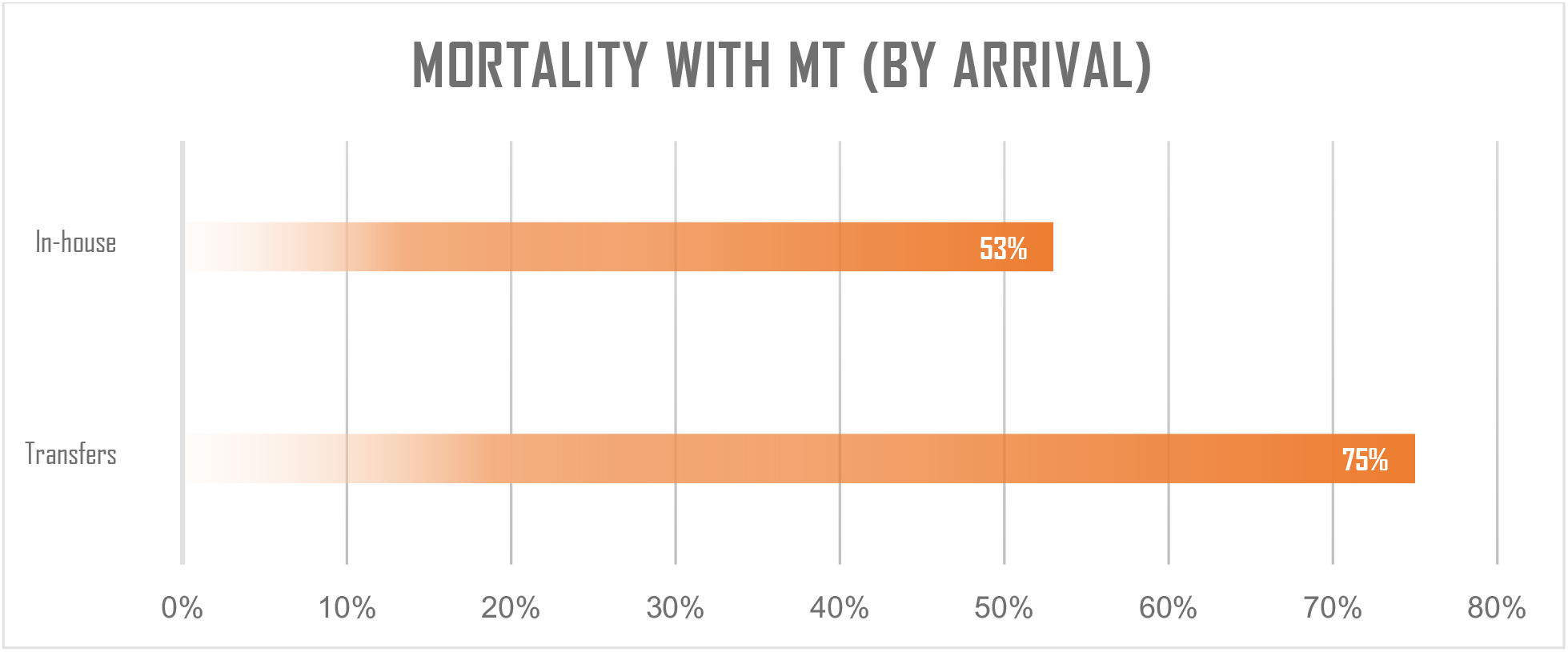
Comparing in-house mortality and transfers. Mortality rate in patients receiving mechanical thrombectomy (MT) by initial arrival to the comprehensive stroke center vs. inter-facility transfer

## DISCUSSION

When taken together, the overall outcomes were poor with high morbidity and mortality in this age group following intervention for acute stroke. MT did not appear to improve outcomes over IVT alone despite patients having a better initial mRS and successful reperfusion with >TICI 2b. Transfers in particular faced worse mortality rates as compared to in-house patients, though this was not a significant difference. When comparing outcomes to other retrospective studies, *Agarwal et al. note* only 44% of nonagenarian patients in their study were discharged to rehab, which is even lower than seen in our cohort. ^[5]^

A lack of delayed outcome data, such as 90-day mRS and disposition, is a notable limitation of this study. The source data is also lacking on baseline comorbidities, however, low sample size makes it difficult to control for these regardless, as it is for other interventional variables that were available such as time from onset to thrombolysis/revascularization.

Multiple factors can be speculated as contributing to poor outcomes in the very elderly. First, they are more prone to develop systemic complications such as infections, particularly aspiration pneumonia in the setting of ischemic stroke ^[9]^. Additionally, elderly brains also have a lower tolerance to ischemia, with infarction progression despite MT, and variably higher sICH rates.^[10]^

While withholding treatment based on age alone is not an ethical (nor a viable) course, these results reinforce the importance of setting realistic expectations of outcomes after AIS in this age group, with informed discussions between physicians and family members when offering interventional management to make the best patient-centered decision. Long-distance transfers for MT often require the deployment of intensive resources, incurring both high costs and potentially delaying their availability for other patients with often equally high acuity issues. In addition, families are often heavily burdened with needing to travel that same distance to visit/stay with the patient while admitted.

Finally, it should not be overlooked that the landmark thrombectomy studies ^[1][2]^ excluded patients with baseline mRS of 3 or higher, though, in practice, it is still frequently offered. In our population, there were examples of MT performed on patients even with a mRS of 5. Without individualized data, it is difficult to say with certainty why this occurred, but from anecdotal experience, it was frequently due to missing baseline history. Due to the high potential harm of delaying stroke intervention, IVT and/or MT are often given as standard of care prior to discovering baseline. This is potentially an easy target to improve resource utilization, particularly in the case of inter-facility transfer, where extra effort to accurately obtain baseline functional status could potentially prevent transfer for futile intervention in an already moribund patient.

A larger scale, multicentered study is necessary to gather more data on nonagenarian outcomes nationwide so evidence-based guidelines can be presented to clinicians and families to guide decision-making when seeking to perform stroke intervention optimally in this special patient population.

## Data Availability

The deidentified data that support the findings of this study are available from the corresponding author, RC, upon reasonable request.

## Acknowledgements

None

